# The effectiveness of multifactorial and multicomponent interventions for the prevention of falls for adults in hospital settings: a systematic review and meta-analysis

**DOI:** 10.1101/2022.05.31.22275666

**Authors:** A.V. Pavlova, P.A. Swinton, L. Greig, L. Alexander, K. Cooper

## Abstract

**Objective:** The objective of this systematic review and meta-analysis was to evaluate the effectiveness of multicomponent and multifactorial interventions for reducing falls in adult in-patients.

**Introduction:** Falls are the most common cause of accidental injury in hospitals worldwide, resulting in high human and economic costs. In attempts to reduce the number of falls, a wide range of interventions have been employed, often in combination, either as a package (multicomponent) or tailored to the individual (multifactorial). There is a need to synthesise the findings from primary studies and assess which approach may be more effective.

**Inclusion criteria:** The systematic review included studies comprising adult inpatients aged 18 years and over from any hospital setting including elective, non-elective, day-case and secondary care. Randomized controlled trials (RCT), cluster-randomised trials, quasi-experimental controlled trials and historical controlled trials were included that presented sufficient information regarding the rate or number of falls.

**Methods:** This effectiveness review was conducted in accordance with JBI methodology and was guided by an a priori protocol. A comprehensive 3-step search strategy was employed across 14 databases. Screening was conducted by two independent reviewers, and data was extracted using a bespoke data extraction tool designed for this review. Methodological quality was assessed using adapted versions of JBI critical appraisal checklists. Meta-analyses were conducted within a Bayesian framework to interpret results probabilistically and account for covariance in multiple sets of falls data reported in the same study. Effect sizes were calculated by comparing the rate or number of falls in the intervention group compared with usual care. Narrative syntheses were conducted on studies that met the inclusion criteria but did not provide sufficient data for inclusion in meta-analyses.

**Results:** A total of 9,637 records were obtained and following screening 24 studies were included in this review, 21 of which presented sufficient information to be included in meta-analyses. Most studies (n=16) comprised a weaker historical control design with 6 quasi-experimental and only 5 RCT studies. Multifactorial interventions were more common (n=18) than multicomponent (n=6), with the most frequent components including environmental adaptations and assistive aids (75% of studies). Meta-analyses provided evidence that both intervention types were effective at reducing the rate and risk of falls compared to usual care. Evidence was also obtained of greater reductions in rate and risk of falls with multicomponent interventions, however, analyses were potentially confounded by an association between intervention type and study design.

**Conclusions:** Falls interventions routinely employed in hospitals can substantially reduce falls, however, no evidence was obtained in support of tailoring interventions to individual risk factors. Future high-quality RCTs are required that directly compare multicomponent and multifactorial interventions.

**Key Points:**

- We found multifactorial and multicomponent interventions to be effective at reducing hospital falls compared to usual care.
- Evidence was obtained that multicomponent interventions were most effective at reducing the risk and rate of falls in hospitals. However, multicomponent interventions were associated with lower quality study designs.
- We found no additional benefit of tailoring intervention components based on an individual’s fall risk factors.
- There is a need for high quality randomised controlled trials comparing multifactorial and multicomponent interventions in hospitals.

## Background and Objectives

Falls in hospital settings are common and comprise events that can lead to serious injury and reduce patient independence. An international consensus statement defined a fall as “an unexpected event in which the participant comes to rest on the ground, floor or lower level” [1]. Individuals over the age of 65 are most affected and falls are responsible for approximately 646,000 deaths a year [2]. The incidence of falls in hospitals range from 6.45 to 18 falls per 1,000 occupied bed days in Australian and UK hospitals [3] [4]. Associated with high rates of falls are substantial costs to individuals and health services [5]. Given their potential consequence, substantial effort and resources are used to limit in-hospital falls as much as possible [6]. An extensive range of interventions have been employed to reduce falls including the use of fall detection devices, environment adaptions, staffing approaches, education of patients and medication/pharmaceutical approaches [7] [8].

Fall prevention interventions can involve a single component (such as education) or a combination of multiple components (e.g. education as well as rounding and environmental adaptations). In a recent scoping review of health technologies used for fall prevention and detection the majority of records (178 out of 411) reported on interventions with multiple components [9]. Multiple component interventions have been reported as either ‘multifactorial’ or ‘multicomponent’. Multifactorial interventions include an initial fall risk assessment which then tailors the intervention components to be used based on individual risk factors. Multicomponent interventions, on the other hand, involve the same bundle of interventions being provided to all patients regardless of fall risk. There currently appears to be no consensus on which approach is more effective at preventing or reducing in-hospital falls.

Avanecean and colleagues [10] conducted a systematic review of the effectiveness of patient centred interventions for fall prevention. However, they focused on the acute hospital setting (medical and surgical units), their search was conducted in 2016, and they were unable to conduct a meta-analysis at the time. Due to the body of literature on multicomponent and multifactorial interventions [9], and the lack of consensus on which is most effective, we identified the need for an updated systematic review comparing the effectiveness of multifactorial and multicomponent interventions for fall prevention in the hospital setting in order to inform this important area of practice. A search of the JBI Database of Systematic Reviews and Implementation Reports, Cochrane Central Register of Controlled Trials (CENTRAL), Medline, CINAHL and PROSPERO identified that no systematic reviews have been published, or are currently underway, on this topic.

## Research Design and Methods

### Review Objective and Questions

The aim of this review was to evaluate and compare the effectiveness of multicomponent and multifactorial interventions employed in hospitals to reduce falls. The specific objectives of this review were: 1) to evaluate the effectiveness of multicomponent or multifactorial interventions in comparison to usual care in prevention of falls in hospital settings; and 2) to identify the most effective combination of interventions for the prevention of falls in hospital settings.

### Protocol and registration

This systematic review was conducted in accordance with JBI methodology for systematic reviews of effectiveness [11] and followed an a priori protocol registered in the PROSPERO database (registration number CRD42019143208).

### Inclusion Criteria

#### Types of studies

This review considered randomized controlled trials (RCT), cluster-randomised trials, quasi-experimental controlled trials and historical controlled trials for inclusion.

#### Participants

This review included studies investigating falls in adult (aged 18+) in-patients, defined as being admitted for patient care activity within any hospital setting including elective, non-elective (emergency admission/Accident & Emergency), day-case and secondary care (community hospital). Studies investigating interventions for fall prevention that included adults in the community, nursing homes or supported living environments were excluded.

#### Types of interventions

Studies investigating either multicomponent or multifactorial intervention strategies to reduce falls in adult hospital patients were included. Multicomponent strategies were defined as interventions where the same bundle of health intervention components were given to all participants [1] [12]. In contrast, multifactorial strategies were defined as interventions where the bundle of health intervention components differed between participants and were based on an assessment of falls risk. The specific features of the multicomponent or multifactorial interventions were further subcategorised using the Prevention of Falls Network Europe (ProFANE) taxonomy [13] (Table 1).

**Table 1.**
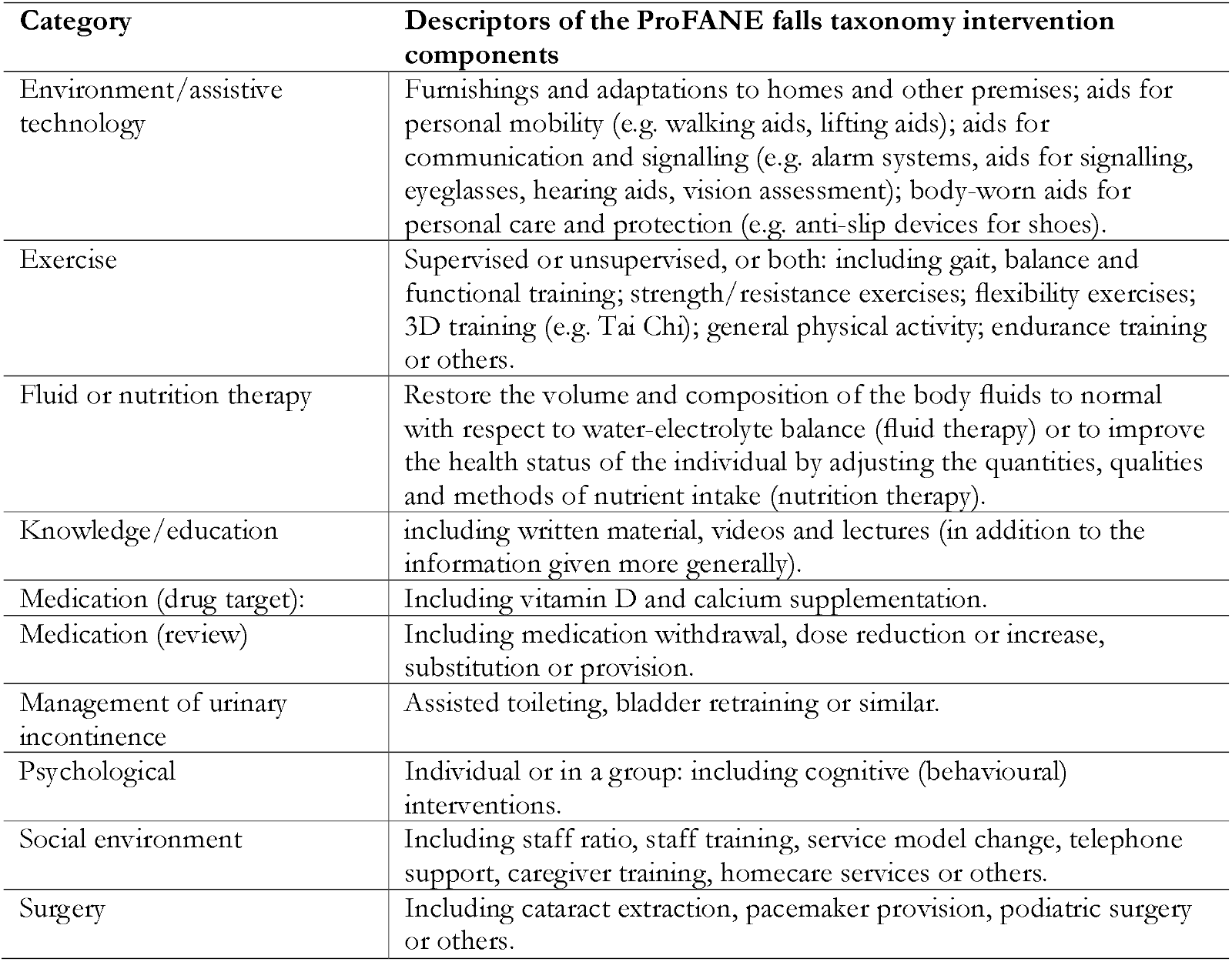
ProFANE (Prevention of Falls Network Europe) taxonomy for fall intervention components.

| Category | Descriptors of the ProFANE falls taxonomy intervention components |
| --- | --- |
| Environment/ assistive technology | Furnishings and adaptations to homes and other premises; aids for personal mobility (e.g. walking aids, lifting aids); aids for communication and signalling (e.g. alarm systems, aids for signalling, eyeglasses, hearing aids, vision assessment); body-worn aids for personal care and protection (e.g. anti-slip devices for shoes). |
| Exercise | Supervised or unsupervised, or both: including gait, balance and functional training; strength/ resistance exercises; flexibility exercises; 3D training (e.g. Tai Chi); general physical activity; endurance training or others. |
| Fluid or nutrition therapy | Restore the volume and composition of the body fluids to normal with respect to water-electrolyte balance (fluid therapy) or to improve the health status of the individual by adjusting the quantities, qualities and methods of nutrient intake (nutrition therapy). |
| Knowledge/education | including written material, videos and lectures (in addition to the information given more generally). |
| Medication (drug target): | Including vitamin D and calcium supplementation. |
| Medication (review) | Including medication withdrawal, dose reduction or increase, substitution or provision. |
| Management of urinary incontinence | Assisted toileting, bladder retraining or similar. |
| Psychological | Individual or in a group: including cognitive (behavioural) interventions. |
| Social environment | Including staff ratio, staff training, service model change, telephone support, caregiver training, homecare services or others. |
| Surgery | Including cataract extraction, pacemaker provision, podiatric surgery or others. |

#### Types of outcome measures

Studies that reported raw data or statistics related to rate or number of falls were included. Studies that reported only specific types of fall (e.g. injurious falls) were excluded as well as those that focused on intermediate outcomes and did not report falls specifically as an outcome.

### Search strategy

The search strategy for this review was part of a larger scoping review investigating general approaches to adult falls prevention and detection in hospital settings [9]. The search strategy was developed in consultation with a research librarian, with the aim of identifying published and unpublished literature. JBI methodology was followed and included a three-step search strategy. An initial limited search was performed in Medline and CINAHL (EBSCOhost) followed by an analysis of the text words contained in the resulting titles, abstracts, keywords and index terms used to describe the publications. A search strategy tailored to each information source based on the identified keywords and index terms was subsequently developed and executed. Finally, the reference lists of all included studies were hand searched for additional sources.

The following databases were searched for published literature: Medline, Joanna Briggs Institute of Systematic Reviews and Implementation Reports, CINAHL, AMED, EmBASE, PEDro, Epistimonikos, EPPI-Centre (DoPHER and TRoPHI), Cochrane Library (controlled trials and systematic reviews), ACM Digital, Compendex, IEEE Xplore, and Science Direct. Grey literature was identified through the following: Google Scholar, Ethos, Mednar, OpenGrey. Trial registries that were searched included: Clinicaltrials.gov, ISRCTN Registry, The Research Registry, European Union Clinical Trials Registry (EU-CTR), and Australia New Zealand Clinical Trials Registry (ANZCTR). In addition, government health department websites and websites of professional bodies were searched for information relating to fall prevention and detection. An example detailed search strategy is presented in Supplementary Files (SF1). Only studies written in the English language were included and to ensure relevance, searches were limited from 2009 to October 2019.

### Study screening and selection

Following the exclusion of duplicates, the titles and abstracts of all studies were screened independently by two reviewers and assessed for their relevance to the review based on the information contained therein. Full-text manuscripts were then retrieved for those studies that met the inclusion criteria. Where reviewers were uncertain about a study’s relevance, these were included at this screening stage and the full-text was also retrieved. Full-text screening of each study was performed independently by two reviewers applying the inclusion and exclusion criteria. Any disagreements between reviewers were resolved by discussion or by a third reviewer where necessary. Articles identified via reference lists were assessed for relevance based on their title and abstracts and those meeting inclusion criteria added to the full-text screening stage. During full-text screening for the larger scoping review [9], relevant studies were identified as being suitable for inclusion in this effectiveness review.

### Data extraction

Prior to data extraction an extraction tool was piloted and discussed within the research team. The tool was then edited to best inform the review questions (SF2). Data extraction was performed by individual reviewers and included the following information where available: title, authors, publication year, journal, country of origin, aims/purpose, study type, intervention categorisation (multifactorial vs multicomponent), intervention structure (inclusion of elements based on ProFANE categorisation), population, sample, setting, outcomes, rate ratio, rate ratio confidence interval, number of fallers in treatment, number of patients in treatment, number of fallers in control and number of patients in control.

### Methodological quality

Adapted versions of JBI’s critical appraisal checklists for RCTs, quasi-experimental and cohort designs were used to assess the methodological quality of included studies [11]. The checklists assessed selection, performance, attrition, detection and reporting bias. Methodological quality of all included studies were assessed independently by two reviewers with discussion used to resolve conflicts.

### Data analysis

Meta-analyses of intervention effects were conducted separately with rate and number of falls data. Rate of falls was taken as the total number of falls per unit of person time and was generally reported in studies as a rate ratio (RaR) between intervention and control groups. The logarithm of the rate ratio was used to pool data and the within-study standard error calculated using the relevant confidence interval provided. Where both adjusted and unadjusted rate ratios were reported, only unadjusted values were selected for analysis. Where rate of falls with confidence intervals were reported separately for treatment and control groups, Monte Carlo simulation (n=10,000) was used to generate RaRs with quantiles of the log transformed sample used to calculate the treatment effect and standard error. For number of fallers the treatment effect was established by calculating a risk ratio (RR) comparing the number of people who fell once or more relative to the total number of individuals admitted to the ward in the treatment group in comparison to the same calculation performed with the control group.

Where data were presented across multiple time points, an intervention effect was calculated at each point and each effect included in the pooled analysis. All meta-analyses were conducted within a Bayesian framework to enable intuitive interpretation of results through reporting subjective probabilities rather than null hypothesis tests or frequentist confidence intervals [14]. Meta-analyses included three-level hierarchical models to account for random variation across studies and covariance between multiple outcomes reported from the same study (e.g. multiple testing points). Small-study effects (publication bias, etc.) were visually inspected with funnel plots and quantified with a multi-level extension of Egger’s regression with effect sizes regressed on within-study variances and weights obtained from the reciprocal of the within- and between-study variances [15] [16].Inferences from all analyses were performed on posterior samples generated by Hamiltonian Markov Chain Monte Carlo with the median (0.5-quantile) and 95% credible intervals (CrIs) constructed to enable probabilistic interpretations of central parameter values and 75% CrIs for variance parameters. Analyses were performed using the R wrapper package brms that interfaced with Stan to perform sampling [17]. Convergence of parameter estimates was obtained for all models with Gelman-Rubin R-hat values below 1.1 [18].

## Results

As part of the larger scoping review a total of 13,553 records were identified through database searching with a further 586 from other sources. Following the exclusion of 4,502 duplicates and 8,845 records that were deemed irrelevant in relation to the inclusion criteria a total of 792 studies were identified for full text examination. Following full text screening, 768 studies were excluded (including 397 as a result of non-inclusion or insufficient reporting of a multicomponent or multifactorial intervention). Of the 24 studies included in this review (Table 2 and SF3 for reference list), 21 provided sufficient data to be incorporated into the meta-analysis (Figure 1).

**Table 2.**
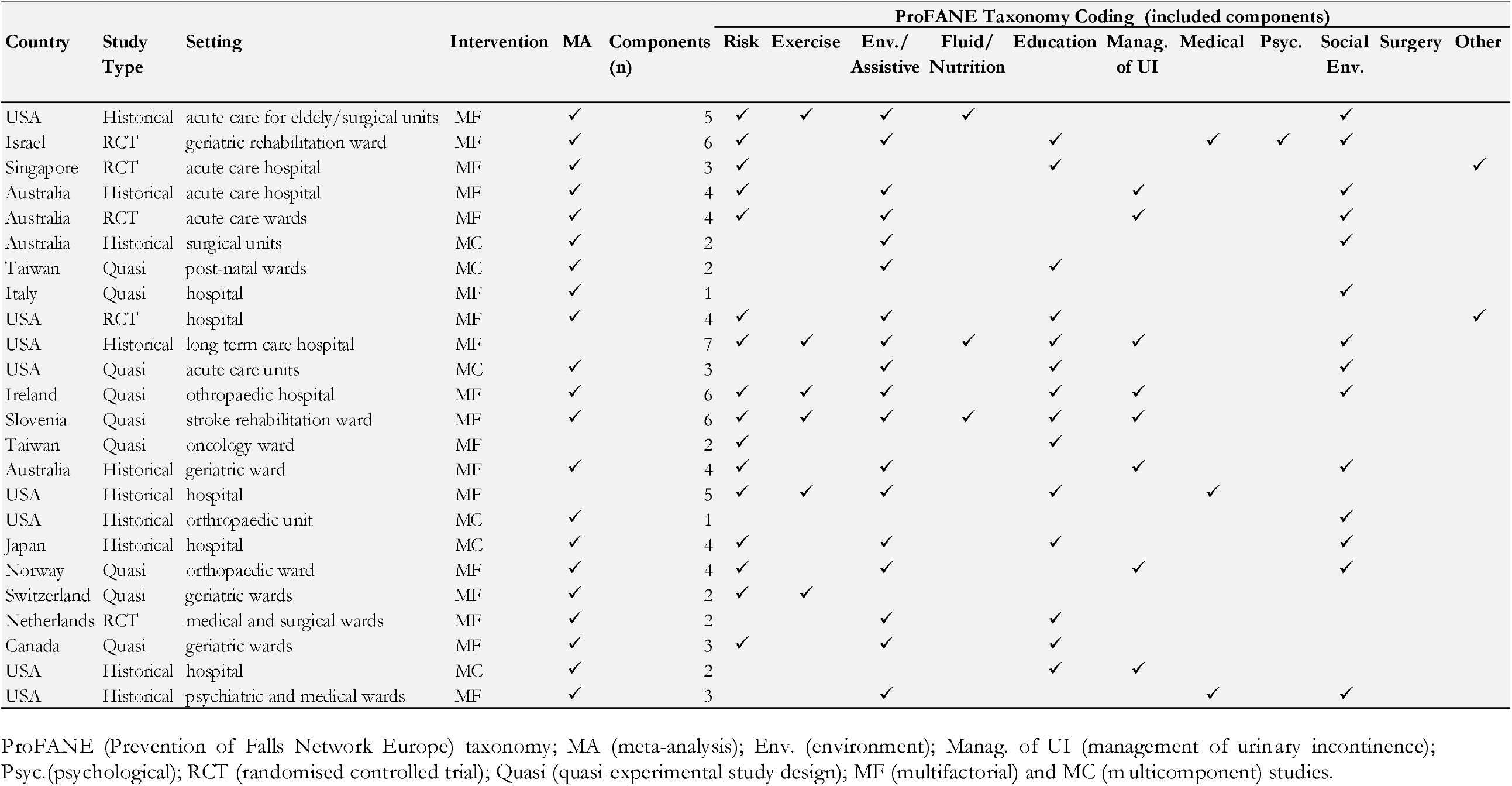
Table of included multifactorial (MF) and multicomponent (MC) studies.

**Figure 1.**
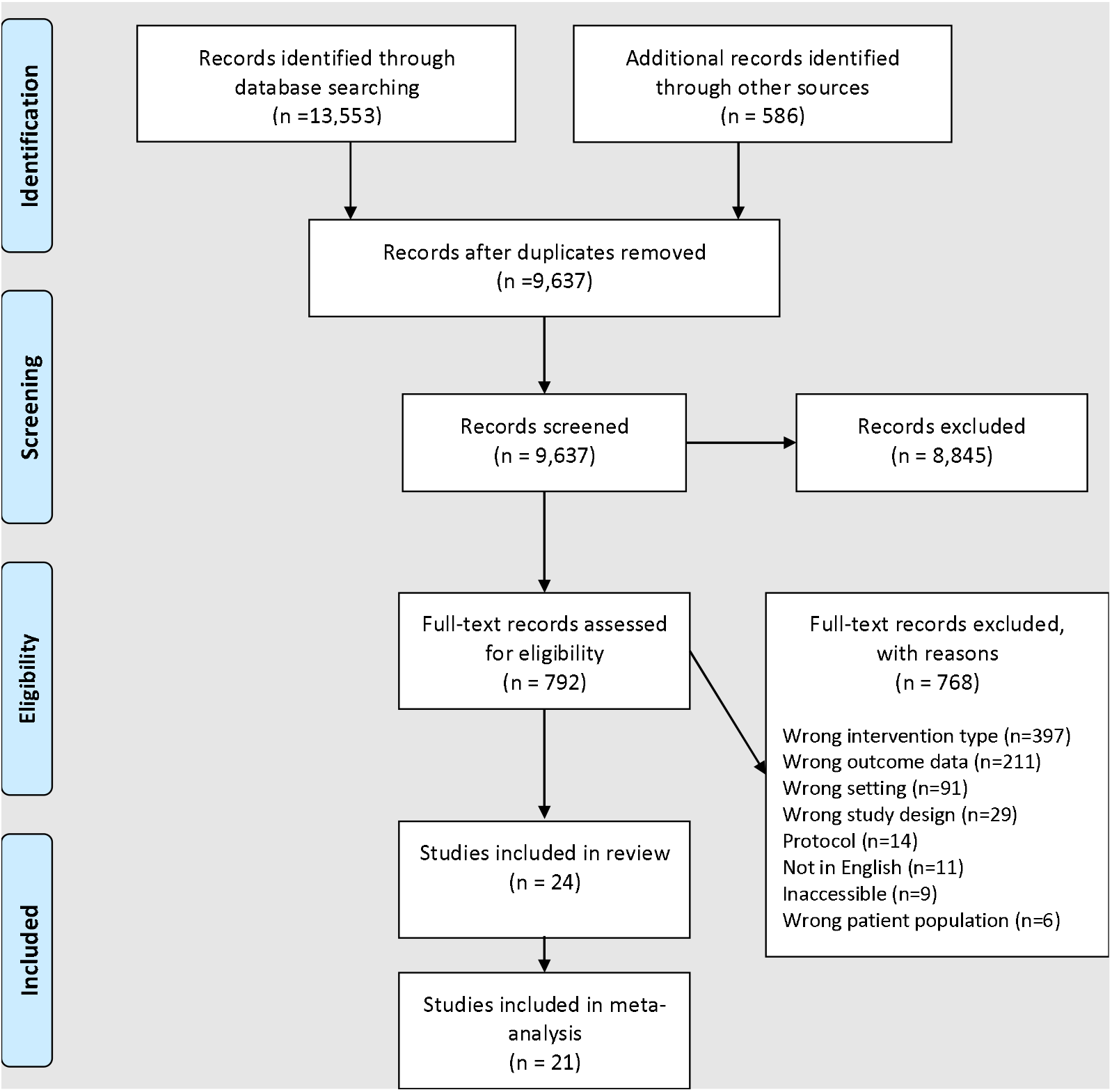
PRISMA Flow diagram showing the number of records at each stage of the literature search and screening process, including reasons for exclusion. From: Page MJ, McKenzie JE, Bossuyt PM, Boutron I, Hoffmann TC, Mulrow CD, et al. The PRISMA 2020 statement: an updated guideline for reporting systematic reviews. BMJ 2021;372:n71. doi: 10.1136/bmj.n71

### Included Studies and Study design

The included studies were conducted across 12 countries (Australia = 4; Canada = 1; Ireland = 1; Israel = 1; Italy = 1; Japan = 1; Netherlands = 1; Norway = 1; Singapore = 1; Slovenia = 1; Switzerland = 1; Taiwan = 2; USA = 8) and comprised three different study designs (historical control = 13; quasi-experimental = 6; RCT = 5).

### Participants and Settings

All included studies reported on hospital patients in a variety of settings including acute care [19] [20] [21] [3] [22], geriatric wards [23] [24] [25] [26], surgical units [27], orthopaedic wards or hospital [28] [29] [30], stroke rehabilitation ward [31], post-natal ward [32], psychiatric in-patient ward [33], long term care [34], oncology [35] and general hospital wards [36] [37] [38] [39] [40] [41].

### Interventions

The most common intervention type was multifactorial, with 18 studies reporting on this type of intervention and the remaining 6 studies reporting a multicomponent intervention. The number of intervention components included across all studies ranged from two to seven, with 8 studies (33%) including two components, 4 studies (17%) including three components, 6 studies (25%) including four components, 2 studies (8%) including five components, 3 studies (13%) including six components, and 1 study including seven components (4%). The most commonly included component was Environment/assistive interventions which were incorporated in 75% of studies and involved actions such as adaptations to the hospital space, aids for personal mobility (e.g. walking aids, lifting aids) and a range of communication and signalling systems. Distribution of intervention components across the included studies is presented in Figure 2.

**Figure 2.**
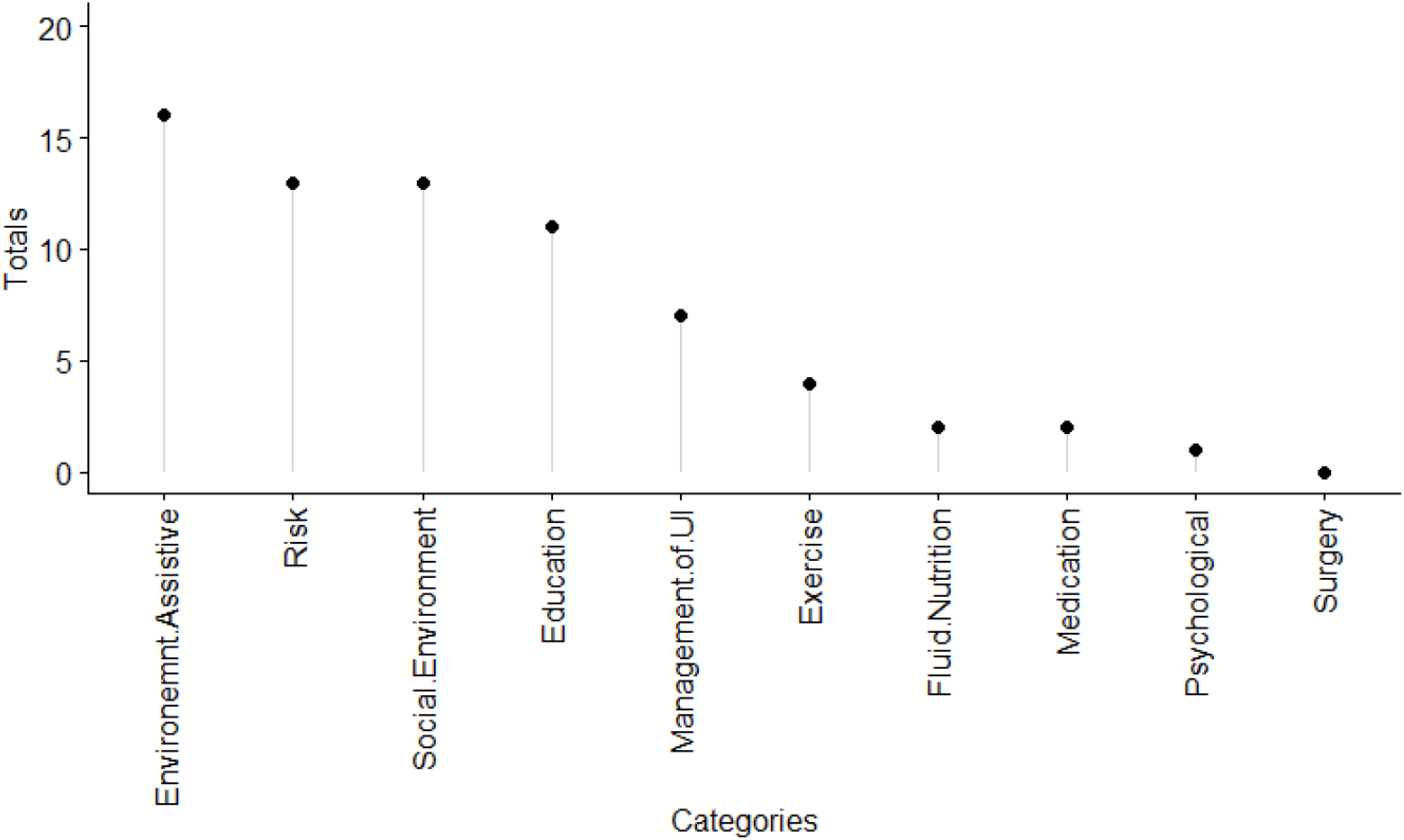
Distribution of intervention components across included studies, showing the total in each category of the ProFANE (Prevention of Falls Network Europe) taxonomy. UI = urinary incontinence.

### Methodological Quality

Methodological appraisal of the five RCT studies are presented in table 3. High scores were achieved for all elements assessed except for blinding (participants, those delivering interventions, and outcome assessors), which would be challenging given the context of the intervention and assessment of falls from central records. Methodological appraisal of the six quasi-experimental studies is presented in table 4. Relatively low scores were obtained for collection of multiple measurements pre- and post-intervention, similarity of patients across conditions, and similarity of treatment beyond intervention. Low scores in the latter category were primarily due to limited reporting of treatment for the control condition. Methodological appraisal of the 13 historical controlled studies is presented in table 5. Low scores were obtained for similarity of patients across conditions and strategies (including statistical) to account for confounders in the final analysis.

**Table 3.** Critical appraisal of studies comprising randomized controlled design

| Study | Q1 | Q2 | Q3 | Q4 | Q5 | Q6 | Q7 | Q8 | Q9 | Q10 | Q11 | Q12 | Score |
| --- | --- | --- | --- | --- | --- | --- | --- | --- | --- | --- | --- | --- | --- |
| Aizen 2015 | Y | U | U | N | N | N | Y | Y | Y | U | Y | Y | 6/12 |
| Ang 2011 | Y | Y | Y | Y | N | N | Y | Y | Y | Y | Y | Y | 10/12 |
| Barker 2016 | Y | Y | Y | N | N | N | Y | Y | Y | Y | Y | Y | 9/12 |
| Dykes 2010 | U | U | Y | U | U | U | Y | Y | Y | Y | Y | N | 6/12 |
| van Gaal 2011 | N | U | Y | N | N | N | U | Y | Y | Y | Y | N | 5/12 |
| % | 75 | 50 | 75 | 25 | 0 | 0 | 75 | 100 | 100 | 75 | 100 | 75 |  |

Q1: True randomization; Q2: Allocation concealment; Q3: Similar baseline; Q4: Participant blinding; Q5: Treatment deliverers blinding; Q6: Outcome assessors blinding; Q7: Similarity beyond treatment; Q8: Participants analysed in original group; Q9: Consistency in outcomes measured; Q10 Reliability of outcomes measured; Q11: Appropriate statistical analysis; Q12 Appropriate trial design

**Table 4.** Critical appraisal of studies comprising quasi-experimental design

| Study | Q1 | Q2 | Q3 | Q4 | Q5 | Q6 | Q7 | Q8 | Score |
| --- | --- | --- | --- | --- | --- | --- | --- | --- | --- |
| Dal Molin 2018 | Y | N | Y | N | N | Y | Y | Y | 5/8 |
| Chen 2010 | Y | Y | U | Y | Y | Y | Y | Y | 7/8 |
| France 2017 | Y | U | Y | N | Y | Y | Y | Y | 6/8 |
| Royset 2019 | Y | Y | U | Y | N | Y | Y | Y | 6/8 |
| Trombetti 2013 | Y | Y | U | Y | N | Y | Y | Y | 6/8 |
| Vieira 2013 | Y | Y | Y | Y | N | Y | Y | Y | 7/8 |
| % | 100 | 67 | 50 | 67 | 33 | 100 | 100 | 100 |  |

Q1: Clear cause and effect; Q2: Similar participants in comparison; Q3: Similarity beyond treatment; Q4: Inclusion of control group; Q5: Multiple measurements pre and post; Q6: Consistency of outcomes measured; Q7 Reliability of outcomes measured; Q8: Appropriate statistical analysis.

**Table 5.** Critical Appraisal of studies comprising historical control design

| Study | Q1 | Q2 | Q3 | Q4 | Q5 | Q6 | Score |
| --- | --- | --- | --- | --- | --- | --- | --- |
| Abdalla 2017 | N | Y | Y | Y | Y | Y | 5/6 |
| Barker 2009 | Y | Y | Y | Y | Y | Y | 6/6 |
| Burston 2015 | Y | U | U | Y | Y | Y | 4/6 |
| Eckstrom 2016 | U | N | N | Y | N | Y | 2/6 |
| Galbraith 2011 | N | N | N | Y | Y | Y | 3/6 |
| Goljar 2016 | U | N | N | Y | Y | Y | 3/6 |
| Huang 2015 | U | N | N | Y | N | Y | 2/6 |
| Isaac 2018 | U | N | N | Y | Y | Y | 3/6 |
| Johnson 2011 | U | N | N | Y | Y | N | 2/6 |
| Lohse 2012 | N | N | N | Y | Y | Y | 3/6 |
| Ohde 2012 | Y | N | N | Y | Y | Y | 4/6 |
| Walsh 2018 | U | N | N | Y | Y | Y | 3/6 |
| Yates 2012 | Y | N | N | Y | N | Y | 3/6 |
| % | 31 | 15 | 15 | 100 | 77 | 92 |  |

Q1: Similar participants in comparison; Q2: Confounders identified; Q3: Strategies to deal with confounders; Q4: Reliability of outcomes measured; Q5: Sufficient follow up time; Q6: Appropriate statistical analysis.

### Meta-analysis Outcomes

Sixteen of the 21 included studies presented sufficient data to calculate rate ratios (RaR), generating a total of 32 intervention effects to be meta-analysed. Similarly, 16 of the included studies presented data to calculate risk ratios (RR), generating a total of 29 intervention effects to be meta-analysed.

#### Rate of falls

Pooling of all 32 effect sizes (multicomponent = 7, multifactorial =25) provided evidence that rate of falls could be reduced through intervention. The Bayesian random effects model (Figure 3) produced a RaR_0.5_ intercept estimate of 0.81 [95%CrI: 0.63 to 1.02] with considerable statistical heterogeneity ([ln τ]_0.5_: 0.26 [75%CrI: 0.18 to 0.34]) and moderate within-study covariance (ICC_0.5_: 0.37 [75%CrI:0.32 to 0.43]). The probability that the effect estimate favoured intervention was equal to *p* = 0.967. The addition of intervention type as a covariate provided evidence of a greater intervention effect with multicomponent compared with a multifactorial intervention. Specifically, the effect of multicomponent interventions was estimated to produce a RaR_0.5_ intercept of 0.68 [95%CrI: 0.47 to 0.98] with the multifactorial effect estimated to produce a RaR_0.5_ intercept of 0.94 [95%CrI: 0.72 to 1.18]. The probability that rate of falls was reduced more in multicomponent compared with multifactorial was equal to *p* = 0.926. The estimated difference between intervention types, however, was confounded with study design, as all studies comprising multicomponent interventions included lower quality quasi-experimental or historical case-controlled designs. Small study effects were also indicated by Egger’s multilevel regression test ([ln *β*_0_]_0.5_: -0.33 [95%CrI: -0.63 to -0.07], average inverse log standard error =9.6).

**Figure 3.**
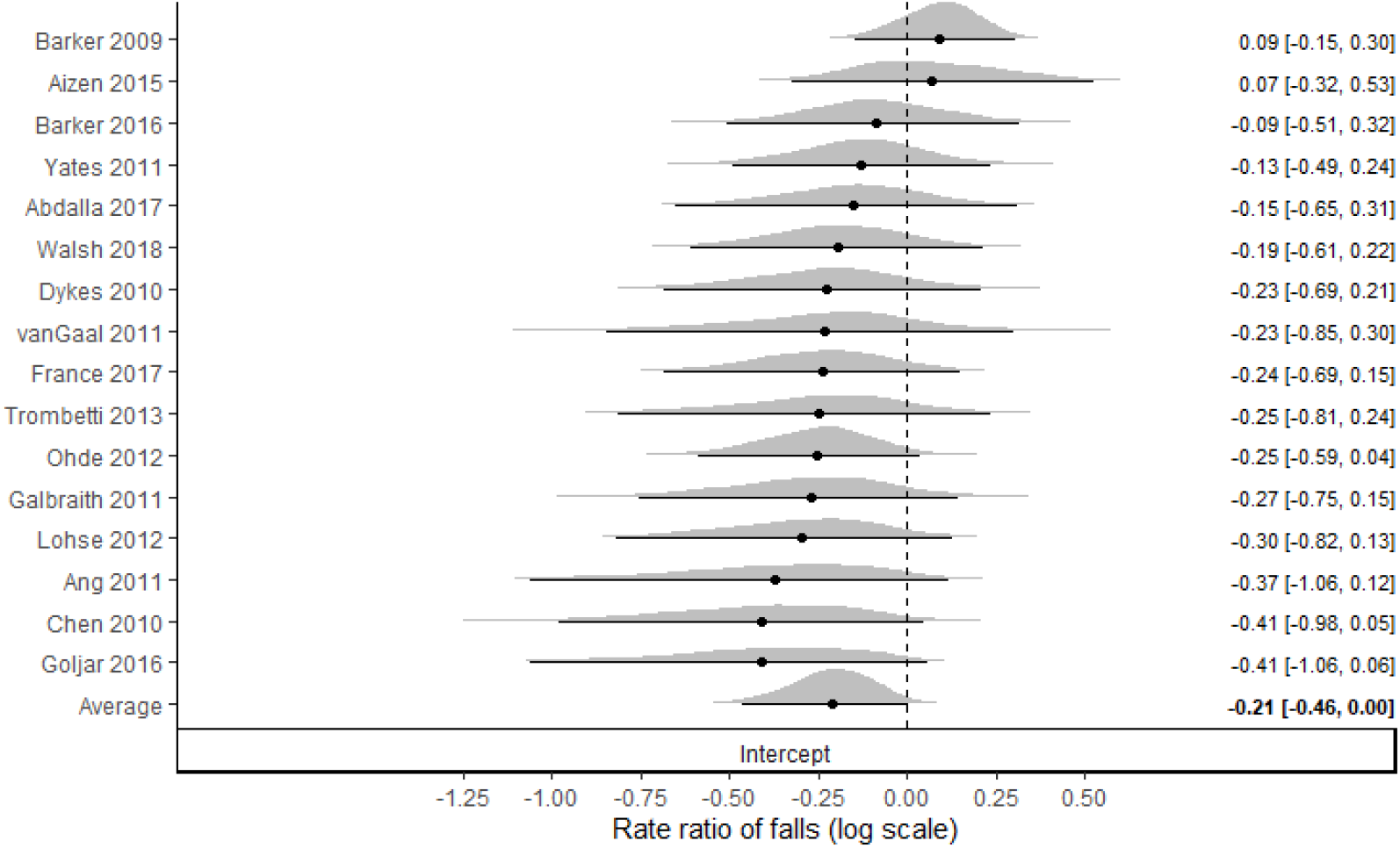
Bayesian forest plot of rate of falls for both multicomponent and multifactorial interventions. Values less than 0 on the log scale represent a decrease in rate of falls in intervention compared to control. Distributions represent “shrunken estimates” based on all effect sizes obtained from the study, the random effects model fitted and borrowing of information across studies to reduce uncertainty. Black circles and connected intervals represent the median value and 95% credible intervals for the shrunken estimates.

#### Risk of falls

Similar results to those obtained for rate of falls were also found for risk of falls. Whilst the median and right tail estimate were shifted to values indicating reduced effectiveness, the pooled results provided evidence for a reduction in risk of falls with intervention. Pooling of all 29 effect sizes (multicomponent = 7, multifactorial =22) with a multilevel Bayesian random effects model (Figure 4) produced a RR_0.5_ intercept estimate of 0.81 [95%CrI:0.61 to 1.05] with considerable statistical heterogeneity ([ln τ]_0.5_: 0.43 [75%CrI: 0.36 to 0.53]) and limited within-study covariance (ICC: 0.18 [75%CrI:0.15 to 0.22]). The probability that the effect estimate favoured intervention was equal to *p* = 0.922. The addition of intervention type as a covariate provided evidence of a greater intervention effect with multicomponent compared with a multifactorial intervention. Specifically, the intervention effect of multicomponent interventions was estimated to produce a RR_0.5_ intercept of 0.62 [95%CrI: 0.37 to 1.03] with the multifactorial intervention effect estimated to produce a RR_0.5_ intercept of 0.90 [95%CrI: 0.65 to 1.21]. The probability that rate of falls was reduced more in multicomponent compared with multifactorial was equal to *p* = 0.891. As noted for rate of falls, the same confounding was present for risk of falls with all multicomponent interventions being conducted within lower quality quasi-experimental or historical case-controlled designs. No substantive evidence of small study effects were identified via Egger’s multilevel regression test ([ln *β*_0_]_0.5_: -0.20 [95%CrI: -0.53- to 0.09], average inverse log standard error = 9.6).

**Figure 4.**
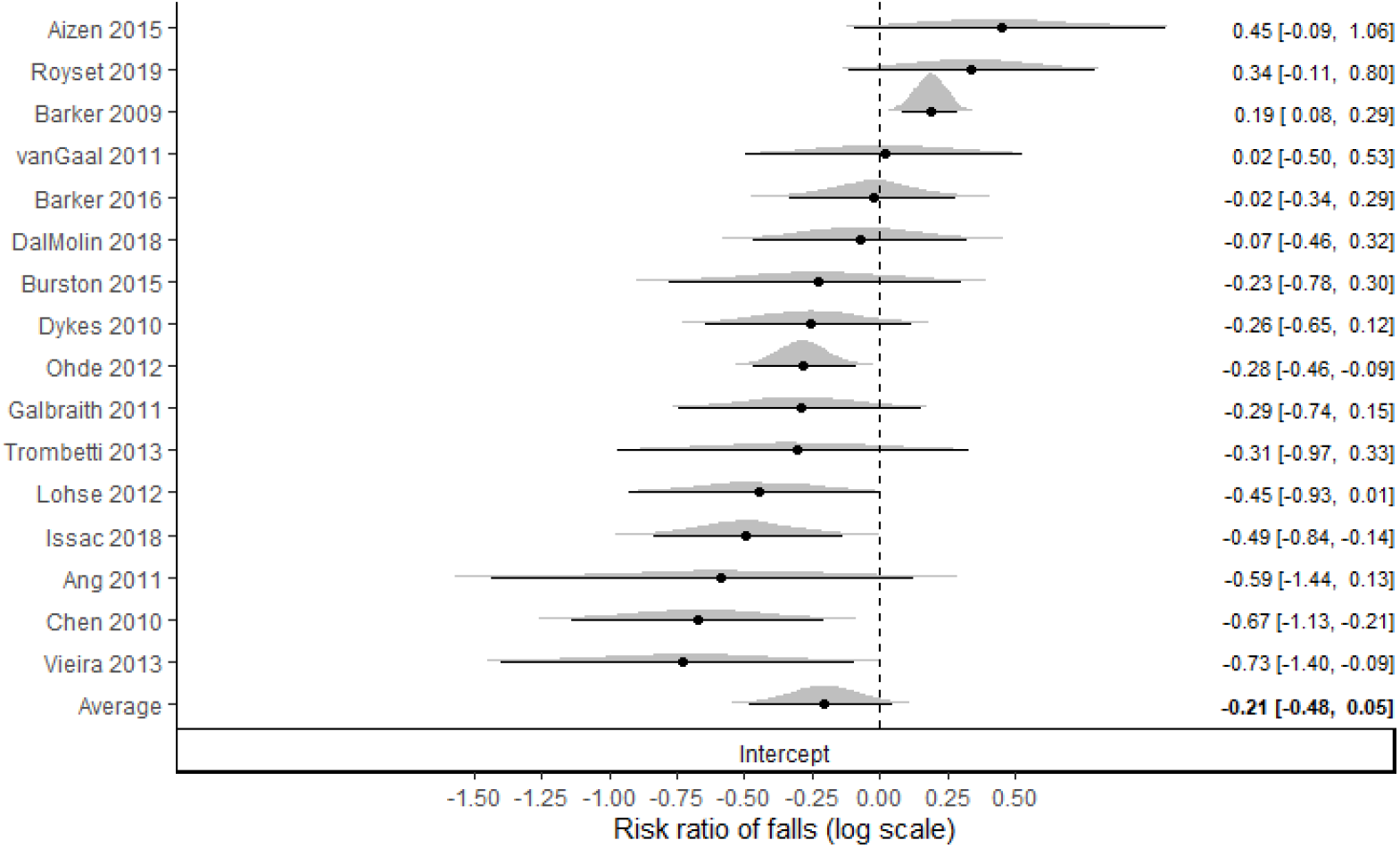
Bayesian forest plot of risk of falls for both multicomponent and multifactorial interventions. Values less than 0 on the log scale represent a decrease in risk of falls in intervention compared to control. Distributions represent “shrunken estimates” based on all effect sizes obtained from the study, the random effects model fitted and borrowing of information across studies to reduce uncertainty. Black circles and connected intervals represent the median value and 95% credible intervals for the shrunken estimates.

### Result summary of studies not included in meta-analysis

Three studies met the inclusion criteria but did not include sufficient data to be incorporated into the meta-analysis [35] [34] [40]. Each of the three studies incorporated a multifactorial intervention and were conducted using a weaker historical controlled design. The findings generated across the studies were mixed with limited ability to synthesise the information due to contrasting results, different populations and interventions, and substantive differences in the number of falls experienced. Eckstrom et al. [34] delivered five intervention workshops and “coaching” for implementation throughout a single year. The workshop focused on evidence-based strategies to decrease the risk of falls for those in long-term care by including screening for falls; assessing gait, balance, orthostatics and other medical conditions; exercise including tai chi; vitamin D supplementation; medication review and reduction; and environmental assessment. The rate of falls (12.9% vs 12.2%) did not change significantly during the three-month post-intervention data collection period compared to the three-month pre-intervention period. In contrast, significant reductions in falls were reported by Johnson et al. [40] with an intervention focussing on falls education for patients and nurses. Across the three-year period which included baseline assessment and intervention, the yearly fall rate reported by Johnson et al. [40] in a general hospital ward decreased from 3.5 to 2.3%. Finally, Huang et al. [35] investigated the effects of a patient centred fall prevention program where patients actively participated in their own safety management after being educated in a range of fall risk scenarios. Comparison data from historical controls were obtained over a three-month period and data collected from the intervention group over a one-year period from the same oncology unit. Limited falls data were obtained, with three falls reported over the historical period and zero falls reported over the intervention period.

## Discussion

Hospital based falls remain a global issue and there is an urgent need for effective intervention programmes. This systematic review followed on from a larger scoping review, funded by the UK National Health Service, into health technologies used for the prevention and detection of falls in hospitals [9]. From the initial scoping review, it was identified that there was a need to perform an effectiveness review to compare multicomponent and multifactorial interventions. The results from the meta-analyses provide clear evidence that interventions in general had the potential to reduce the rate and risk of adult falls in hospitals. However, large between study variance highlights that effectiveness may be influenced by a range of factors including the research design, the intervention (e.g. variations in the risk assessment used and the type and number of components), the patient characteristics and ward settings (e.g. vast differences between geriatric, post-natal or general wards). Considering the large emphasis on falls in hospitals globally [42] [43] [44] [45] [46] the number of primary studies formally investigating the effectiveness of interventions on falls is limited. Only 21 studies met the criteria to be included in this review, whereas our previous scoping review identified 404 articles relevant to falls in hospital. In addition, most of the studies included in this review were limited to quasi-experimental or historical control studies [9] with only five RCTs identified.

Contrary to the popular opinion that multifactorial interventions are superior due to tailoring of interventions to the falls risk of an individual, no evidence was obtained to support this position. Indeed, the findings of the meta-analyses provided strong statistical support for the superiority of multicomponent interventions to reduce the rate and risk of falls. However, these findings are likely to be confounded with research quality, whereby all multicomponent studies were restricted to lower quality quasi-experimental and historical controlled trials. Due to this likely confounding, coupled with substantive heterogeneity and potential small-study effects (identified for rate of falls), it is difficult to make a clear recommendation based on the evidence obtained here. However, given the additional resources and time required by staff to conduct fall risk assessments and individually tailor intervention components, based on the evidence obtained here this strategy may not be the most efficient. In an already busy and potentially short-staffed hospital environment managers may need to consider the balance between the time and staff required to perform risk assessments compared to giving everyone the same intervention.

Additional heterogeneity was identified in multifactorial studies where a range of approaches were used to assess falls risk and allocate individual health technologies. Other factors may also influence study findings. Avancean and colleagues [10] identified that three multifactorial RCT studies, out of the five included in their review, had a significant reduction in fall rates. They attributed length of stay (and thus intervention), lack of communication between staff, and adherence to fall prevention programmes as potential barriers to success in the two other studies. Staff non-compliance with interventions can also be an issue [47] [48] and is a frequently reported barrier to intervention success; however, very few of the included studies assessed or reported on this aspect.

The methodological quality of each study was assessed for selection, performance, attrition, detection and reporting bias using JBI critical appraisal tools appropriate for each design. Only five of the studies included here were RCTs and many of the quasi-experimental studies scored low on important methodological points. Most studies followed a weak design using historical controls where subsequent changes in falls risk may be due to a range of factors beyond the intervention allocated. In addition, many of the historical controlled trials were over very short time periods, for example Yates and Tart [33] compared a year of post-implementation data with just 5 months baseline fall data, with an intervention involving medication review, non-slip socks, and staff and patient education. Eckstrom and colleagues [34] reported falls data between baseline and intervention periods which were just 3 months each, and intervention effects may have been confounded by implementation compliance. In contrast, some studies were collected over extended periods. Barker and colleagues [21] assessed their multifactorial targeted intervention, including environmental adaptations, signage, supervision and bed/chair alarm, over a 9-year period. Walsh and colleagues reported on a series of multicomponent interventions over an 11-year period while Ohde and colleagues [38] assessed their 4-component intervention over a 6-year period. However, it is important to consider that over these longer time periods the general culture of fall prevention may have changed (e.g. changes in policies and practice or uptake) and so it is challenging to account for the variation in falls due to specific interventions versus more global factors. Furthermore, a large proportion of the historical designs did not control for potential confounders in patient characteristics between samples.

The findings of this effectiveness review must be interpreted in the context of its limitations. There was a substantive imbalance between studies investigating multifactorial (n=18) versus multicomponent (n=6) interventions, highlighting the general consensus that multifactorial interventions are likely to be superior. Additionally, the included studies may not represent the breadth of health technologies available, with most focusing on manipulating environment design and its specific sub-components. As a result of the relatively small study numbers and focus on environmental manipulations we were unable to address our second objective to identify the most effective intervention components. Collectively, this leads us to make some recommendations for future research. Given the potentially surprising finding of superior results with multicomponent interventions, future RCTs should directly compare multicomponent and multifactorial interventions. These studies should be supplemented with economic and resource data to determine not only effectiveness in falls but associated costs. Additionally, future studies should be clear regarding their definitions of multifactorial and multicomponent interventions, which was used, and should categorise intervention components according to the ProFANE falls taxonomy.

In summary, the findings of the present review identify no benefit to tailoring interventions based on individual risk assessments, although the presence of confounding is likely. These findings are in accord with recent advice that fall risk assessments are discouraged and fall prevention interventions should be provided to all. The National Institute for Health and Care Excellence (NICE) updated their recommendations in 2013 to advise against the use of fall risk prediction tools (those that class individuals as ‘at risk/not at risk’ or class individuals into ‘low/med/high risk’ of falling) and instead called for all individuals 65 years and over (and those 50-64 with an underlying condition) to be regarded as at risk of falling [49].

## Supporting information

Supplement file PRISMA checklist

## Data Availability

All data produced in the present study are available upon reasonable request to the authors

## Acknowledgments

We would like to thank Colin Maclean, research librarian, for support with literature searching. We thank Dr Clare Bostock, consultant geriatrician, and Rosie Cooper, falls lead and improvement advisor, for their expert input to the study steering committee. Thank you to the service users for their input at steering committee meetings.

## Supplementary Files

### SF1. Search strategy and number of hits at each stage of the search

SF1. Search strategy and number of hits at each stage of the search.
| Source | Search | Hits |
| --- | --- | --- |
| Medline | 1. TX Hospital* AND MH (Accidental falls) OR TX (“fall* prevention” OR “fall* detection”)<br>2. MH (Delivery of health care OR biomedical technology) OR TX (Technolog* OR device* OR intervention* OR strateg* OR system* OR organiz* OR organis* OR program*)<br>3. 1 AND 2 | 1. 8,184<br>2. 8,112,408<br>3. **2,558** (limits applied) |
| JBISRIIR | 1. Hospital* AND fall* | 1. **283** |
| CINAHL | 1. TX Hospital*<br>2. MH (Accidental falls) OR TX (“fall* prevention” OR “fall* detection”)<br>3. MH (Biomedical enhancement OR health care delivery) OR TX (technolog* OR device* OR intervention* OR strateg* OR program* system* OR organiz OR organis*)<br>4. 1 AND 2 AND 3 | 1. 1,005, 248<br>2. 15, 984<br>3. 1,710,131<br>4. **2,482** (limits applied) |
| AMED | 1. Hospital*<br>2. Accidental falls OR (“fall* prevention” OR “fall* detection”)<br>3. (Biomedical technology OR delivery of health care) OR TX (technolog* OR device* OR intervention* OR strateg* OR system* OR organiz* OR organis*)<br>4. 1 AND 2 AND 3 | 1. 12,967<br>2. 2,167<br>3. 75, 481<br>4. **111** (limits applied) |
| EMBASE | 1. Hospital*<br>2. Accidental falls OR “fall* prevention” OR “fall* detection”<br>3. 1 AND 2 | 1. 7,199,688<br>2. 5,624<br>3. **2,414** |
| PEDro | 1. Fall* AND hospital* | 1. **240** |
| Epistimonikos | 1. Hospital<br>2. Fall* prevention OR Fall* detection<br>3. 1 AND 2 | 1. 72,088<br>2. 37,066<br>3. **1219** (1503) |
| DoPHER | 1. Fall* AND hospital* | 1. **14** |
| TRoPHI | 1. Fall* AND hospital* | 1. **38** |
| Cochrane | 1. Fall* AND hospital* | 1. **126** |
| ACM Digital | 1. Hospital*<br>2. Fall prevention<br>3. Hospital AND fall prevention | 1. 2,432<br>2. 13,724<br>3. **60** |
| Compendex | 1. Fall prevention<br>2. Hospital<br>3. 1 AND 2 | 1. 7,523<br>2. 19,1301<br>3. **359** |
| IEEE Xplore | 1. Hospital*<br>2. Accidental falls OR “fall* prevention” OR “fall* detection”<br>3. 1 AND 2 | 1. 82, 259<br>2. 1,518<br>3. **129** |
| Science direct | 1. Hospital<br>2. Technology<br>3. “fall prevention” OR “fall detection”<br>4. 1 AND 2 AND 3 | 1. 3,123,153<br>2. 3,781,010<br>3. 3,744<br>4. **2,293** |

Search performed in October 2019

### SF2. Data extraction tool adapted for use in falls effectiveness review

**SF2. Data extraction tool adapted for use in falls effectiveness review.**
| **Field** | **Instructions/Description** |
| --- | --- |
| Study | Author year (e.g. Smith 2010) |
| Title | Title of article |
| Journal | Journal title |
| Country of Origin | Country the study took place in |
| Aims/Purpose | Aim/purpose of the study |
| Study Type | Select study type from dropdown list:<br>• Historically controlled trial<br>• Quasi-experimental (non-randomised)<br>• RCT |
| Intervention/Technology Category | Select from drop down list:<br>• Multicomponent<br>• Multifactorial |
| Population | Who did they aim to recruit? Eg. adults, older people, patients, nurses |
| Study Sample | Sample size, age, % females, pathology (if available) of the study sample. |
| Setting | Study setting, eg. hospital or specific ward. |
| Intervention Information | Description of the intervention |
| ProFANE Taxonomy Coding | Tick all intervention categories that apply:<br>• Risk<br>• Exercise<br>• Environment/ Assistive technology<br>• Fluid/Nutrition therapy<br>• Education/knowledge<br>• Management of urinary incontinence<br>• Medication (drug target or review)<br>• Psychological<br>• Social Environment<br>• Surgery<br>• Other (make a note) |
| Outcomes Reported | List all the outcomes they report relating to fall prevention and detection.<br>Example:<br>• Fall number<br>• Rate of falls (e.g. rate per 1000 patient days) |
| Findings/Conclusions/Recommendations | Brief summary of main findings and conclusions or recommendations related to fall prevention and detection. |
| Reviewer Comments | Any additional comments |
| Initials and Date of Extraction | Please initial and date each study row |

